# Assortative mating and parental genetic relatedness drive the pathogenicity of variably expressive variants

**DOI:** 10.1101/2023.05.18.23290169

**Authors:** Corrine Smolen, Matthew Jensen, Lisa Dyer, Lucilla Pizzo, Anastasia Tyryshkina, Deepro Banerjee, Laura Rohan, Emily Huber, Laila El Khattabi, Paolo Prontera, Jean-Hubert Caberg, Anke Van Dijck, Charles Schwartz, Laurence Faivre, Patrick Callier, Anne-Laure Mosca-Boidron, Mathilde Lefebvre, Kate Pope, Penny Snell, Paul J. Lockhart, Lucia Castiglia, Ornella Galesi, Emanuela Avola, Teresa Mattina, Marco Fichera, Giuseppa Maria Luana Mandarà, Maria Grazia Bruccheri, Olivier Pichon, Cedric Le Caignec, Radka Stoeva, Silvestre Cuinat, Sandra Mercier, Claire Bénéteau, Sophie Blesson, Ashley Nordsletten, Dominique Martin-Coignard, Erik Sistermans, R. Frank Kooy, David J. Amor, Corrado Romano, Bertrand Isidor, Jane Juusola, Santhosh Girirajan

## Abstract

We examined more than 38,000 spouse pairs from four neurodevelopmental disease cohorts and the UK Biobank to identify phenotypic and genetic patterns in parents associated with neurodevelopmental disease risk in children. We identified correlations between six phenotypes in parents and children, including correlations of clinical diagnoses such as obsessive-compulsive disorder (R=0.31-0.49, p<0.001), and two measures of sub-clinical autism features in parents affecting several autism severity measures in children, such as bi-parental mean Social Responsiveness Scale (SRS) scores affecting proband SRS scores (regression coefficient=0.11, p=0.003). We further describe patterns of phenotypic and genetic similarity between spouses, where spouses show both within- and cross-disorder correlations for seven neurological and psychiatric phenotypes, including a within-disorder correlation for depression (R=0.25-0.72, p<0.001) and a cross-disorder correlation between schizophrenia and personality disorder (R=0.20-0.57, p<0.001). Further, these spouses with similar phenotypes were significantly correlated for rare variant burden (R=0.07-0.57, p<0.0001). We propose that assortative mating on these features may drive the increases in genetic risk over generations and the appearance of “genetic anticipation” associated with many variably expressive variants. We further identified parental relatedness as a risk factor for neurodevelopmental disorders through its inverse correlations with burden and pathogenicity of rare variants and propose that parental relatedness drives disease risk by increasing genome-wide homozygosity in children (R=0.09-0.30, p<0.001). Our results highlight the utility of assessing parent phenotypes and genotypes in predicting features in children carrying variably expressive variants and counseling families carrying these variants.

## INTRODUCTION

Rare genomic variants, including copy-number variants (CNVs) and single nucleotide variants (SNVs), contribute to a significant proportion of individuals with neurodevelopmental and psychiatric disorders^1–7^. Many of these variants are often inherited and exhibit variable expressivity and phenotypic heterogeneity, in contrast to syndromic variants with invariable clinical presentation^8^. For example, phenotypic heterogeneity has been observed among carriers of rare recurrent SNVs associated with neurodevelopmental disorders^9^, such as the candidate schizophrenia gene *NRXN1*, which also contributes to intellectual disability/developmental delay (ID/DD), autism, and congenital malformations with varying degrees of penetrance^10^. This variable expressivity and phenotypic heterogeneity can be attributed to a multi-hit model, where a primary variant sensitizes the genome for neuropsychiatric features and genetic interactions of the primary variant with other (“secondary”) variants in the genetic background determine the ultimate phenotypic outcome^11–14^. In fact, pathogenic rare CNVs on chromosome 1q21.1, 15q13.3, and 16p12.1 tend to be transmitted from a relatively mildly affected parent, with the increase in phenotypic severity corresponding to an increase in the number (i.e. burden) of secondary variants over generations^11, 14, 15^. These secondary variants are often inherited from the non-carrier parent and have been shown to act synergistically with the primary pathogenic variant^14, 16^. This phenomenon of complex inheritance forms the basis of many neurodevelopmental disorders and complicates predicting the phenotypic trajectory of individuals within families with these disorders.

These complexities necessitate examining other factors, such as parental phenotype and genotype, in affected probands that may elucidate their ultimate phenotypic trajectory. Previous reports have shown that family history is a risk factor for neurodevelopmental disorders, with increased within and cross-disorder risk observed among relatives of individuals with psychiatric disease^17–19^. This increase in liability towards multiple diagnostic outcomes may be due to the overlapping genetic etiologies of neurodevelopmental disorders^20^ and is particularly relevant for understanding mechanisms of variable expressivity of rare variants. Here, we sought to broaden our understanding of the effect of various phenotypic and genetic factors in parents on neurodevelopmental disease outcomes in children. To that end, we examined more than 38,000 spouse pairs and parent pairs of children with neurodevelopmental disorders to identify risk factors that affect disease trajectories in affected children, including clinical and subclinical neurological and psychiatric features and patterns of assortative mating on those features, as well as genetic similarity between parents. We further investigated the implications of these patterns across generations and identified potential mechanisms by which they contribute to disease risk. Our findings highlight assortative mating and parental kinship as previously unexplored risk factors for neurodevelopmental and neuropsychiatric disease.

## SUBJECTS AND METHODS

### Cohort Information

We assessed parental factors affecting neurodevelopmental disease in two North American autism cohorts: the Simons Simplex Collection (SSC)^21^ and the Simons Foundation Powering Autism Research (SPARK)^22^. We analyzed genetic and/or phenotypic data from 2,517 trios from SSC and 7,175 trios from SPARK. We also analyzed genetic and/or phenotypic data from an additional 67 parent pairs in SSC and 1,570 parent pairs in SPARK for families where the relevant proband data were not available.

We also collected genomic and/or phenotypic data for another neurodevelopmental disease cohort of 203 samples from 43 trios, 4 larger families, and 21 spouse pairs carrying the 16p12.1 deletion. Probands in these families were ascertained for developmental delays or other neurodevelopmental phenotypes and were identified as 16p12.1 deletion carriers through clinical diagnostic tests. Families provided informed consent to give blood samples and phenotypic data under Pennsylvania State University Institutional Review Board approved protocol #STUDY00000278, and deidentified data from medical reports were collected under approved protocol #STUDY00017269.

We further assessed assortative mating and spousal relatedness in 26,610 partner pairs from the UK Biobank^23^, identified using Data Return 1872^24^ (Application ID: 45023). UK Biobank is a database containing genetic, lifestyle, and health information from 500,000 volunteer participants from the UK. Additionally, we assessed profiles of genetic relatedness in parent pairs of children ascertained for neurodevelopmental disorders carrying rare copy number variants (CNVs) from another clinical cohort evaluated at GeneDx. This cohort consists of families with affected children referred by clinicians to GeneDx for genetic testing and is distinct from the 16p12.1 deletion cohort. Families were subsequently selected for those carrying pathogenic CNVs (see *CNV calling*), resulting in 357 parent pairs from families carrying 21 distinct disease-associated CNVs (**Supp. Table 1**). The study of this cohort was conducted under an IRB protocol approved by the Western Institutional Review Board (IRB #STUDY 1169768, WIRB Pro Number 20162523), which states that this research meets the requirements for a waiver of consent. We note that the terms “spouses” and “partners” are used interchangeably, while the term “parents” refers to “spouses” or “partners” in the context of affected children in this manuscript.

### DNA Extraction and Sequencing

DNA extraction and sequencing of samples within the SPARK, SSC, and UK Biobank cohorts were performed by the relevant biobank. For directly recruited participants from the 16p12.1 deletion cohort, DNA was extracted from peripheral blood using the QIAamp DNA Blood Maxi extraction kit (Cat. No: 51104, Qiagen), while clinical collaborators sent DNA samples directly to the Girirajan laboratory. SNP arrays for 36 samples were generated as previously described^14^, while 92 additional samples were run on Illumina OmniExpress 24 v 1.1 microarrays at the Northwest Genomics Center at the University of Washington (Seattle, WA, USA). Illumina HiSeq X was used to generate 150-bp paired-end whole genome sequence (WGS) data at ≥30X coverage for 118 individuals by Macrogen Labs (Rockville, MD, USA). DNA extraction and sequencing from the GeneDx cohort were performed as previously described^25^.

### Variant calling

#### CNV calling

CNVs in the GeneDx cohort were first called from exome sequencing data and confirmed via microarray, as previously described^25^. Disease-associated CNVs were identified from any overlap with previously reported breakpoints for pathogenic CNVs^15^.

#### SNV calling

Single nucleotide variants (SNVs) and small indels were identified in 16p12.1 deletion families using the GATK Best Practices pipeline. After removing duplicates with PicardTools^26^, we performed base-pair quality score recalibration and then used GATK HaplotypeCaller v.3.8^27^ to call variants. Calls from all samples were merged to call variants jointly using GATK GenotypeGVCFs v.4.0.11^27^, followed by variant quality score recalibration. Variants were filtered for those with an allele balance between 0.25 and 0.75 or >0.9 and a read depth of ≥8.

Whole genome (for SSC) and whole exome (for SPARK) variant call files (VCFs) were downloaded from SFARI Base (https://base.sfari.org). Whole exome VCFs were obtained from the UK Biobank using Data Field 23157. VCFs from all three cohorts were left-normalized and multi-allelic records were split with BCFtools^28^. Calls were filtered using the same criteria as above. Variants from the 16p12.1 deletion cohort, SSC, and SPARK were additionally filtered for a quality ≥50 and a ratio of quality to alternative allele depth ≥1.5.

For all four cohorts, variant frequency in gnomAD^29^ was annotated using vcfanno^30^ and variants were filtered for those with gnomAD exome and genome frequency ≤0.1%. Variants in the SSC, SPARK, and UK Biobank cohorts were filtered for an intracohort frequency ≤0.1%. To achieve a similar frequency threshold with a smaller sample size, variants in the 16p12.1 deletion cohort were filtered for those that were private within related individuals. ANNOVAR^31^ was used to assign variants to genes from GENCODE v19 (16p12.1 deletion cohort) or GENCODE v38 (SSC, SPARK, and UK Biobank)^32^, and variants in all cohorts were filtered for loss-of-function (LOF), missense, or splice variants in protein-coding genes. LOF variants were those annotated as ’stopgain’, ’stoploss’, ’frameshift_deletion’, ’frameshift_insertion’, or ’frameshift_substitution’ by ANNOVAR, and missense variants were those annotated as ’nonsynonymous_SNV’, ’nonframeshift_deletion’, ’nonframeshift_insertion’, or ’nonframeshift_substitution’. CADD Phred-like scores^33^ were annotated using vcfanno as a deleteriousness metric and missense and splice variants were filtered for those with a CADD Phred score ≥25. For probands within SSC, variants were additionally annotated with SFARI Gene Tier of the affected gene^34^.

#### Polygenic risk score calculations

We obtained autism GWAS summary statistics from Grove and colleagues^35^. Duplicate and ambiguous single nucleotide polymorphisms (SNPs) were removed, and the remaining SNPs were filtered for imputation INFO scores>0.8. We then used PLINK^36^ to perform an initial round of quality control by filtering SNPs with minor allele frequency <0.05, Hardy-Weinberg equilibrium test p-value <1.0×10^-5^, and genotype rate <0.05, and removed samples missing >1% of genotypes or having a Mendelian error rate >5%. We then used the HRC-1000 Genomes Imputation toolkit (https://www.well.ox.ac.uk/#wrayner/tools) to process PLINK files into individual chromosomes for imputation, and used VcfCooker (https://genome.sph.umich.edu/wiki/VcfCooker) to convert PLINK files to VCF files. SNPs were imputed using the TOPMed v.r2 imputation server using Eagle v2.4 for phasing^37^. After imputation, VCF files were converted back to PLINK format, and SNPs were again filtered using the criteria from the initial quality control. SNPs were additionally filtered to remove samples with >±3SD of the mean heterozygosity and strand-flipped to match the GWAS summary statistics. We filtered the GWAS summary statistics for SNPs present in the HapMap3 dataset^38^, and used 1000 Genomes datasets to calculate linkage disequilibrium matrices for the SNPs. After regressing betas of GWAS SNPs according to linkage disequilibrium, we used the LDPred2-auto model^39^ to calculate autism PRS for probands in the SSC cohort.

### Phenotype Assessment

We assessed self-reported psychiatric phenotypes in probands and parents in the SPARK cohort and 23 quantitative autism and cognitive metrics in probands (**Supp. Table 2**) and two quantitative autism metrics in parents (Social Responsiveness Scale (SRS)^40^ and Broad Autism Phenotype Questionnaire (BAPQ)^41^) from the SSC cohort downloaded from SFARI base (https://base.sfari.org). We further assessed neurological and psychiatric phenotypes in 57 parent pairs from 16p12.1 deletion families. Information was collected on each adult family member either through self-reports using a standardized clinical questionnaire or clinician reports. Curated data was generated using a union of clinical diagnoses, self-reported phenotypes, and interpreted responses to clinical questionnaires. We also assessed phenotypic correlations in the UK Biobank using self-reported diagnoses of psychiatric disorders from Data Field 20544.

Linear regression was used to assess the predictive power of parental SRS and BAPQ scores and various genetic factors, including autism PRS and rare CNVs and SNVs, towards child quantitative phenotypes, while tetrachoric correlations were used to assess phenotypic correlations between psychiatric diagnoses in parents and children and between spouse pairs. To minimize the effects of comorbidities and isolate the specific correlations of interest, pairs were removed from cross-disorder tetrachoric correlations if both members of the pair had either diagnosis of interest. For example, when examining the relationship between the female member of the spouse pair having anxiety and their partner having depression, pairs where the male partner had anxiety or the female partner had depression were excluded from the analysis.

### Kinship Calculations

In the 16p12.1 deletion, SSC, SPARK, and UK Biobank cohorts, we followed the same initial SNP quality control steps outlined above for PRS calculations. Additionally, SNPs within long-range linkage disequilibrium regions^42^ were removed, as were samples with ±2 standard deviations of the mean cohort heterozygosity rate. For SPARK, SSC, and the 16p12.1 cohort samples, we next assessed ancestry using fastStructure^43^ to cluster samples (k=5-6) against individuals with known ancestry from the HapMap3 project^38^. Kinship analysis in these cohorts was restricted to pairs with self-reported or calculated European ancestry. UK Biobank samples were filtered for pairs of European ancestry using Data Field 22006.

KING^44^ was used to calculate kinship coefficients for 44 parent pairs from the 16p12.1 cohort, 1,996 parent pairs from the SSC cohort, 4,211 parent pairs from the SPARK cohort, 6,315 partner pairs from the UK Biobank cohort, and 357 parent pairs from the GeneDx cohort. In the GeneDx cohort, 11 parent pairs with a kinship coefficient less than -0.1 were removed from further analysis, as this highly negative value may indicate that these samples come from different ancestral populations.

### Runs of Homozygosity

To identify runs of homozygosity (ROH), we applied the same initial SNP quality control filters described above for PRS calculations on 3,714 probands ascertained for autism in the SPARK cohort and 1,981 probands from the SSC cohort. ROHs were calculated using the PLINK *homozyg* function^36^. We allowed a maximum of one heterozygous and five missing SNPs in each ROH, and runs were restricted to those at least 1 Mbp in length.

### Statistical Analysis

Tetrachoric correlations were performed using the *correlation* package^45^ in R. Linear regression was performed using *statsmodels* in Python, with proband age at the time the Autism Diagnostic Observation Schedule (ADOS) was administered included as a covariate in the models. Pearson correlations and t-tests were performed using the *pearsonr* and *ttest_ind* functions in the *scipy stats* (v. 1.7.1) package^46^ in Python (v. 3.9.7), respectively.

## RESULTS

### Parental phenotypes predict child phenotypes

To understand the relevance of parental phenotypes in neurodevelopmental disorders, we examined the correlation between six parent and child psychiatric phenotypes in the Simons SPARK autism cohort (**Fig. 1A, Table S4A**). We observed significant positive within-disorder correlations between mothers and fathers and male and female probands for most phenotypes (Pearson correlation, R=0.1-0.49, Bonferroni-corrected p<0.05) (**Fig. 1A, Table S4A**). We also observed several cross-disorder correlations, including those common to both sexes, such as the correlation between depression in parents and anxiety in probands (R=0.10-0.27, p<0.01), and those which are unique to one sex, such as the correlation between OCD in fathers and ADHD in female probands (R=0.13, p=0.003), suggesting parental and sex-specific correlations for certain pairs of features (**Fig. 1A, Table S4A**). These correlations recapitulate previous patterns of within- and cross-disorder correlations observed in studies of both psychiatric disease cohorts and cohorts from the general population^17, 47, 48^.

**Figure 1.**
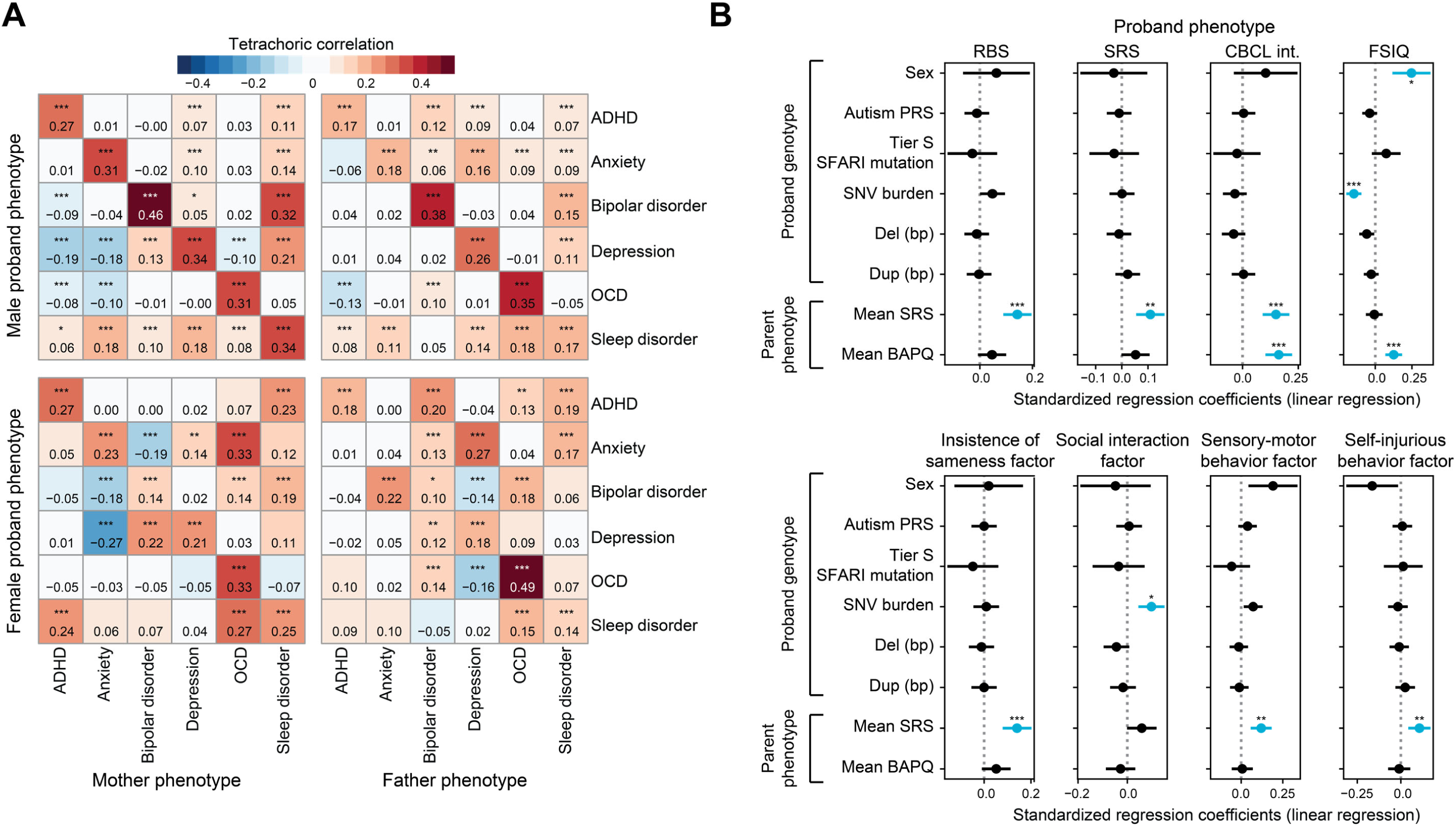
Parental phenotypes predict clinical outcomes in children. (A) Tetrachoric correlations between self-reported psychiatric disorders in mothers and fathers and male and female probands within the SPARK autism cohort (n=592-4,757 for each correlation). (B) Regression coefficients from linear models incorporating effects of both genetic factors and parental phenotypes towards quantitative proband autism and cognitive phenotypes (n=1,628-2,168 for each model). RBS: Repetitive Behaviors Scale, SRS: Social Responsiveness Scale, CBCL int: Child Behavior Checklist 6-18 internalizing, FSIQ: Full-scale IQ. Selected models shown here, full data in Table S4. Blue dots and lines indicate significance after Bonferroni-correction. * Bonferroni-corrected p≤0.05, ** p≤0.01, *** p≤0.001.

To assess the relationship of parent and child phenotypes outside the context of a family history of psychiatric disorders, we used linear models to assess the relationship between 23 quantitative measures of autism and cognitive features (**Table S2**) in probands from the simplex autism cohort SSC and two measures of autism features in their unaffected parents, Social Responsiveness Scale (SRS)^40^ and Broad Autism Phenotype Questionnaire (BAPQ)^41^ scores (**Fig. 1B, Table S4B**). Several measures of autism features in probands were significantly influenced by parental SRS scores, including Repetitive Behaviors Scale (RBS)^49^ (β=0.14, Bonferroni-corrected p=1.95×10^-6^), SRS (β=0.11, p=0.002), and Child Behavior Checklist (CBCL)^50^ internalizing (β=0.15, p=1.11×10^-5^) scores of children with autism were positively associated with biparental mean SRS scores (**Fig. 1B, Table S4B**). Biparental mean SRS and/or BAPQ scores were in fact stronger predictors of autism severity in affected children than any individual group of rare or common genetic variants for several measures (**Fig. 1B, Table S4B**). These data highlight the potential utility of assessing parent phenotypes to predict child phenotypes. Even in simplex families, where parents are largely unaffected, child phenotypes can be predicted by parent phenotypes, suggesting some sub-clinical level of genetic liability in the parents of affected children which compounds within a single generation to surpass a disease threshold. This also indicates the presence of multiple genetic factors which individually are not sufficient to cause overt phenotypes in parents but may additively or synergistically interact in offspring to cause more overt phenotypes in subsequent generations.

### Phenotypic assortment among partner pairs

Given the relevance of parental phenotypes towards predicting phenotypes in affected children, we hypothesized that assortative mating may be a relevant risk factor for neurodevelopmental disease. Assortative mating has been well described for neurological and psychiatric phenotypes^24, 51–58^ and has previously been shown to result in increasing disease liability over generations using national registrar data from Sweden and Denmark^17, 47, 59^. This is akin to patterns of increasing disease liability observed in neurodevelopmental disease cohorts, particularly in families carrying variably expressive variants^11, 14, 15^. Using the 16p12.1 deletion as a paradigm for these disorders, we first examined the frequency of self-reported neurological and psychiatric features in adult family members of children with the 16p12.1 deletion ascertained for neurodevelopmental phenotypes. Adult deletion carriers in our cohort manifested several neurological and psychiatric phenotypes, including seizures (5.8%), schizophrenic features (20%), depression (36%) and anxiety (39%) symptoms, and addiction phenotypes (8.0%) (**Fig. 2A**). In line with expectations of assortative mating, their non-carrier partners also manifested these features at similar frequencies (seizure: 5.8%, schizophrenic features: 12%, depression: 28%, anxiety: 26%, addiction: 10%) (**Fig. 2A**). As these data are indicative of assortative mating, we further assessed spousal assortment based on these phenotypes and were able to identify several significant within- and cross-disorder correlations (**Fig. 2B, Table S4C**). For example, 16p12.1 deletion carriers with anxiety were likely to have partners with anxiety (R=0.69, Bonferroni-corrected p=8.14×10^-7^) or depression (R=0.66, p=0.001) (**Fig. 2B, Table S4C**). These data confirm that the well-documented patterns of assortative mating for psychiatric phenotypes are typically prevalent in families carrying variably expressive variants with a high load of genetic liability for neurodevelopmental disorders. We also compared patterns of phenotypic assortment in parents of children in an affected cohort and the general population by analyzing five self-reported psychiatric phenotypes in the SPARK autism cohort and the UK Biobank (**Fig. 2C-D, Table S4D**). Although these phenotypes occur much more frequently in the SPARK cohort (**Supp. Table 3**), positive within- and cross-disorder correlations were observed in both of these cohorts (**Fig. 2C-D, Table S4D**). For example, women in both cohorts with bipolar disorder or mania were more likely to have partners with bipolar disorder (SPARK: R=0.30, Bonferroni-corrected p=7.69×10^-129^; UKB: R=0.24, p=3.53×10^-304^) or schizophrenia (SPARK: R=0.24, p=3.90×10^-80^; UKB: R=0.50, p=0) (**Fig 2C-D**). This confirms that spousal assortment on psychiatric phenotypes occurs both in individuals ascertained for disease and those from the general population.

**Figure 2.**
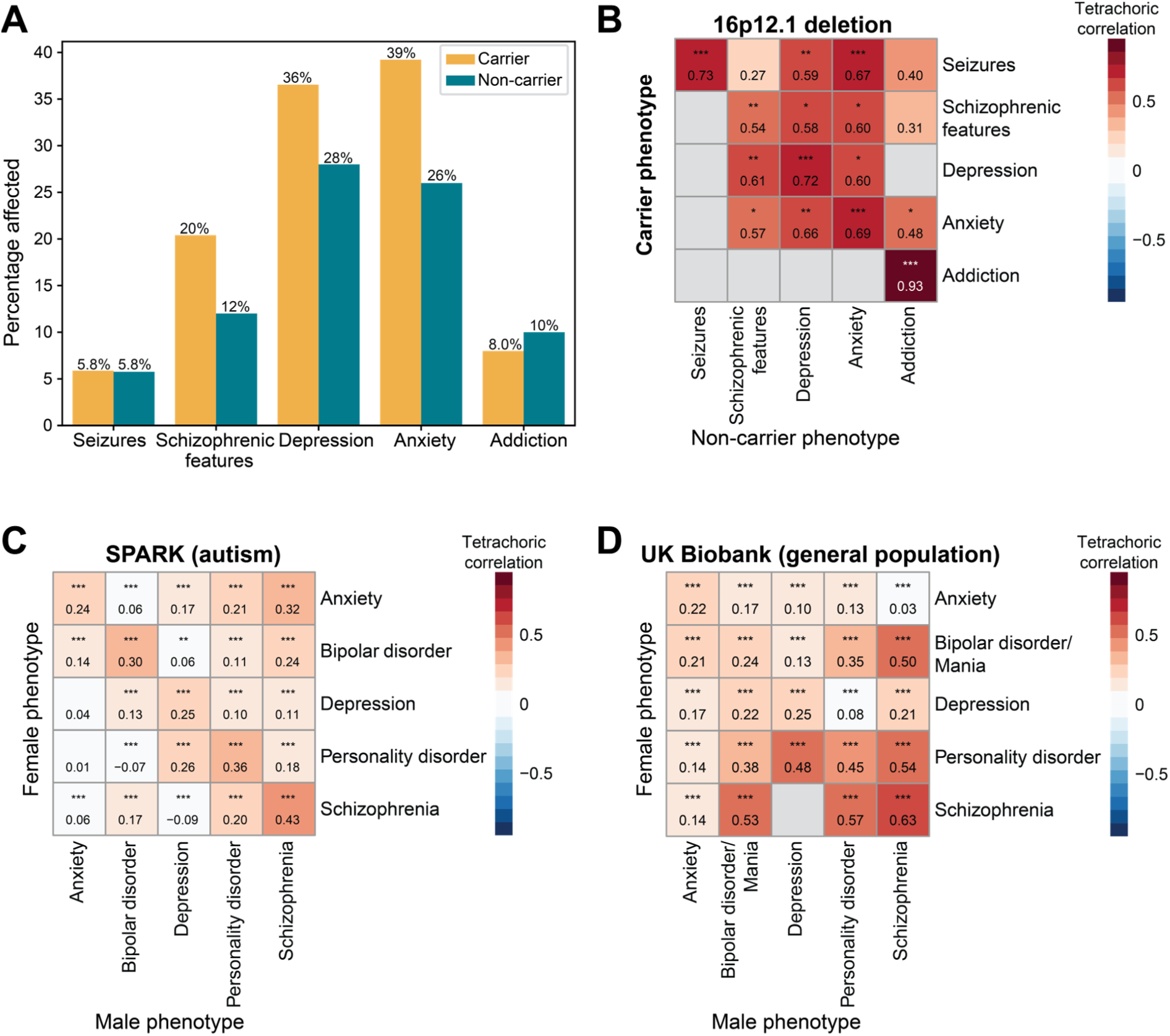
Patterns of assortative mating in multiple cohorts. (A) Percentage of carrier and non-carrier spouses in a cohort of 16p12.1 deletion carrier families with five neurological and psychiatric disorders (n=49-52). (B-D) Tetrachoric correlations of neurological and psychiatric phenotypes between (B) carrier and non-carrier spouses in the 16p12.1 deletion cohort (n=28-50), (C) mothers and fathers in the SPARK autism cohort (n=3,956-6,142), and (D) female and male spousal partners in the UK Biobank (n=19,468-22,953). Grey boxes indicate insufficient sample size for correlation. * Bonferroni-corrected p≤0.05, ** p≤0.01, *** p≤0.001.

### Genetic consequences of assortative mating

Partner assortment will result in positive correlations between their risk alleles, which underly the phenotypes driving assortment^60^. While genetic risk of assortative mating is often measured using association studies of common variants^24, 60–62^, we hypothesized that rare deleterious variants also correlate among partners, given the links between rare variants and psychiatric disorders on which partners tend to assort^7,63, 64^. To confirm this pattern in the assessed cohorts, we correlated the burden of rare deleterious coding single nucleotide variants (SNVs) between partners in each cohort (**Fig. 3A**). We observed that the burden of rare variants positively correlated between partners in the 16p12.1 deletion cohort (Pearson’s R=0.46, p=2.79×10^-4^), SPARK (R=0.52, p<10^-12^), Simons Simplex Collection (SSC, R=0.57, p<10^-12^), and the UK Biobank (R=0.07, p=6.80×10^-9^) cohorts (**Fig. 3A**), confirming that partner assortment based on related phenotypes drives correlations in genetic risk. Interestingly, the correlation was much weaker in the UK Biobank (R=0.07) than the disease cohorts (R≥0.46), possibly due to the lower frequency of psychiatric phenotypes (**Supp. Table 3**) and lower burden of genetic risk in this cohort. The ultimate outcome of this correlation in burden is an increase in the variance of genetic liability over generations^65^, where affected individuals will have more affected children and unaffected or mildly affected individuals will have more mildly affected children (**Fig. 3B**), mimicking patterns observed in families carrying variably expressive variants. For example, 16p12.1 deletion carrier families typically show increases in clinical severity over generations, from unaffected or mildly affected grandparents to parents with psychiatric features and then to children with neurodevelopmental disorders (**Fig. 3C**) and this increase in phenotypic severity corresponds to an increase in genetic burden^11, 14, 16^ (**Supp. Figure 1**). Taken together, this suggests that assortative mating drives the increases in genetic liability that result in the severe phenotypes observed in neurodevelopmental disease cohorts, particularly in carriers of variably expressive variants.

**Figure 3.**
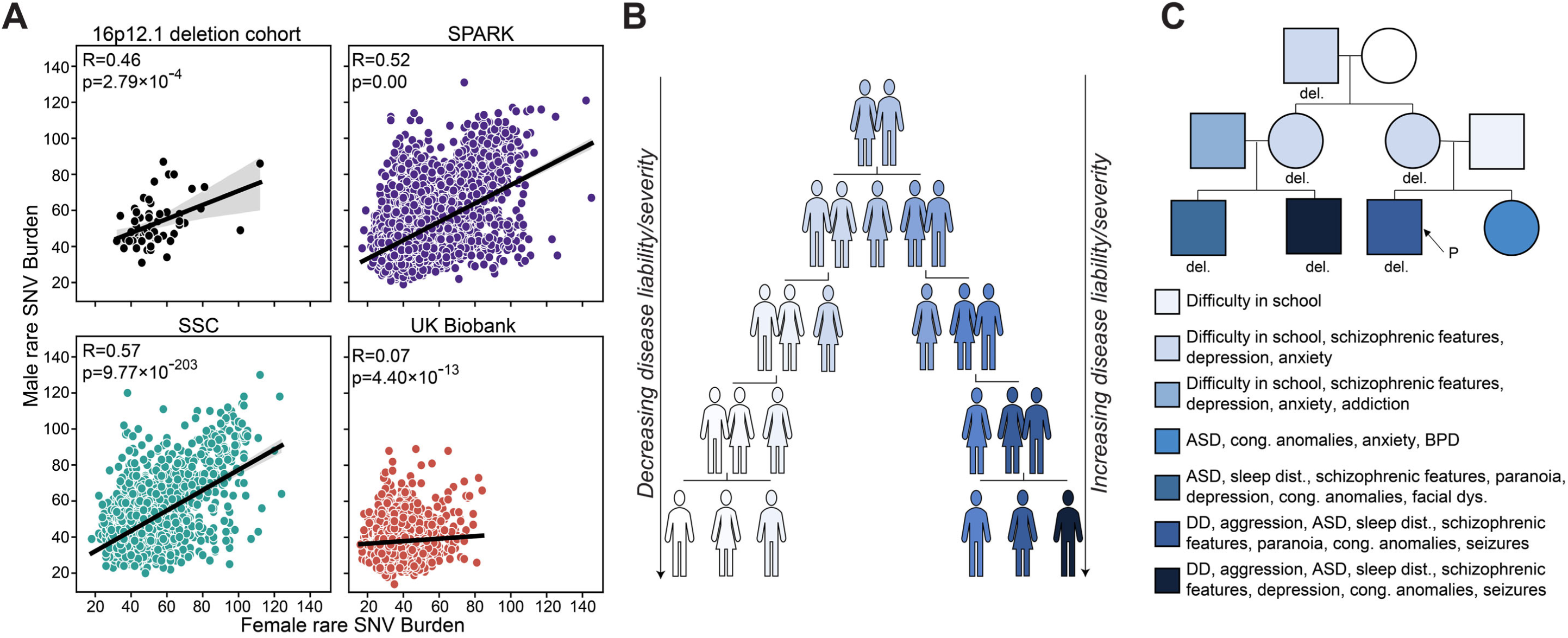
Assortative mating drives increases in genetic risk variance. (A) Pearson correlations of rare, likely deleterious SNVs in female and male partners of spouse pairs from the 16p12.1 deletion cohort (n=59), SPARK (n=7,352) and SSC (n=2,397) autism, and the UK Biobank (n=10,323) cohorts. (B) Schematic illustrating increases in disease liability variance over generations resulting from assortative mating. (C) Example pedigree of assortative mating and compounding disease severity in a 16p12.1 deletion carrier family. White indicates no overt phenotype.

**Figure 4.**
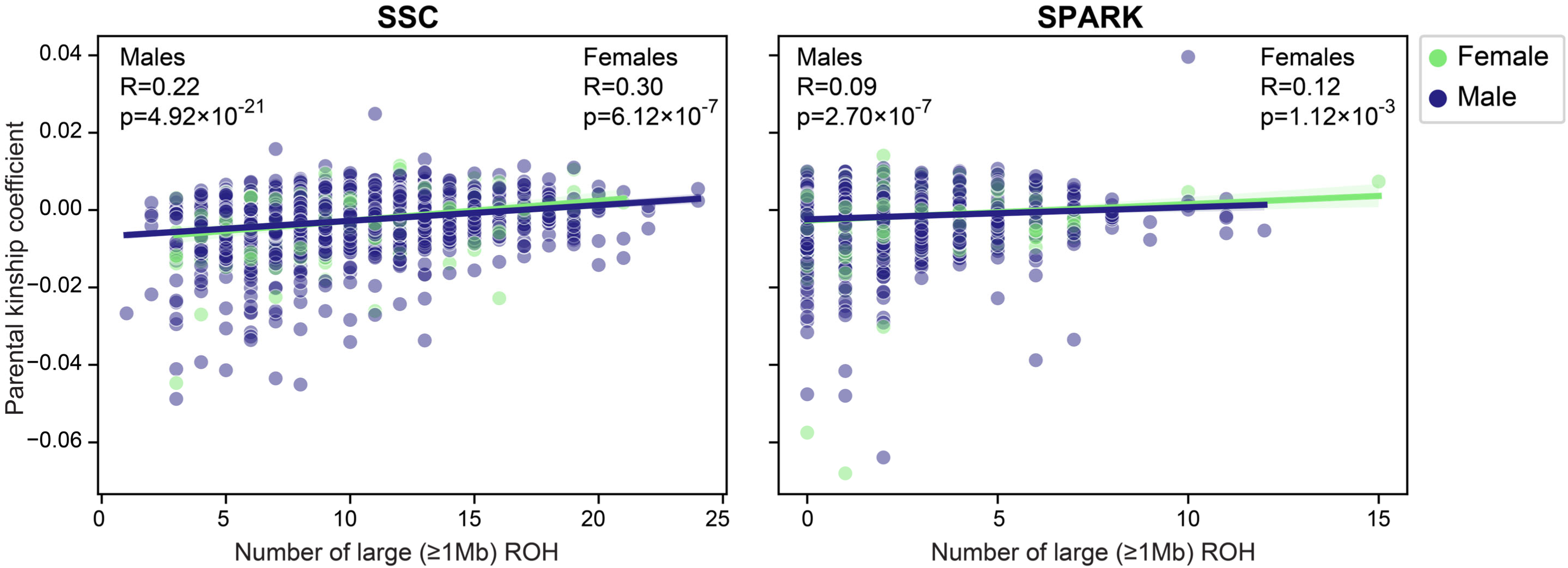
Parental relatedness correlates with homozygosity in children. Pearson correlations of parental relatedness (measured as kinship coefficients) and the number of large (≥1 Mb) runs of homozygosity in male and female probands in the SSC (male n=1,711, female n=270) and SPARK (male n=2,956, female n=758) autism cohorts. probands from both SSC (male R=0.22, p=4.92×10^-21^, female R=0.30, p=6.12×10^-7^) and SPARK (male R=0.09, p=2.70×10^-7^, female R=0.12, p=1.12×10^-3^) (Fig. 4), indicating that increased spousal relatedness could lead to increased disease risk by unmasking recessive alleles within those intervals.

### Parental relatedness as a risk factor for neurodevelopmental disease

Another parental factor which can increase disease risk is the level of consanguinity or genetic relatedness of parents, through increasing both the frequency of autosomal recessive conditions and genome-wide homozygosity^66, 67^. Long stretches of homozygosity have been associated with increased risk for several neurodevelopmental disorders^68, 69^. Although the parents assessed in our cohorts are not consanguineous, we hypothesized that increases in parental relatedness lead to increases in genome-wide homozygosity and increased risk of neurodevelopmental disorders. To assess this, we examined large (≥1 Mbp) runs of homozygosity (ROH) in 3,714 and 1,981 children of the parent pairs analyzed in the SPARK and SSC cohorts, respectively. We found a significant correlation between parental relatedness (as measured by kinship coefficients) and the number of large ROH in both male and female We then asked how this parental relatedness could interact with other neurodevelopmental disease factors. We calculated kinship coefficients as a measure of relatedness between partner pairs from all four cohorts and found negative correlations between spousal kinship and the average spousal rare variant burden in all three neurodevelopmental disease cohorts (16p12.1 deletion: R=-0.30, p=0.04; SPARK: R=-0.13, p=1.05×10^-17^, SSC: R=-0.23, p=1.34×10^-23^) (**Fig. 5A**). However, no significant correlation was observed for partners in the UK Biobank (R=-0.02, p=0.07) (**Fig. 5A**). We then examined parental relatedness in the context of severe damaging mutations in children ascertained for autism in SSC. We found that kinship coefficients of 102 parent pairs of children with loss-of-function mutations in canonical autism-associated genes (Tier S SFARI genes)^34^ were significantly lower than kinship coefficients in 1,251 parent pairs of affected children without damaging mutations in autism-associated genes (one-tailed t-test p=0.041) (**Fig. 5B**). We also examined kinship coefficients of 346 parent pairs from an independent cohort of children ascertained for neurodevelopmental disorders carrying 21 different pathogenic CNVs (**Supp. Table 2**). We found that parental kinship had an inverse relationship with severity as well as penetrance of the primary variant in children. Kinship coefficients were significantly higher among parents of children with inherited CNVs compared to parents of children with *de novo* CNVs (one-tailed t-test, p=0.03) (**Fig. 5C**). Similarly, parents of children with duplications, which are typically associated with incomplete penetrance^70^ and less severe features^71^ than deletions, had higher kinship coefficients compared to parents of children with deletions (one-tailed t-test, p=0.02) (**Fig. 5D**). Although not statistically significant, we also identified a trend of higher kinship values in families with variably expressive CNVs compared to syndromic CNVs (one-tailed t-test, p=0.14) (**Fig. 5E**).

**Figure 5.**
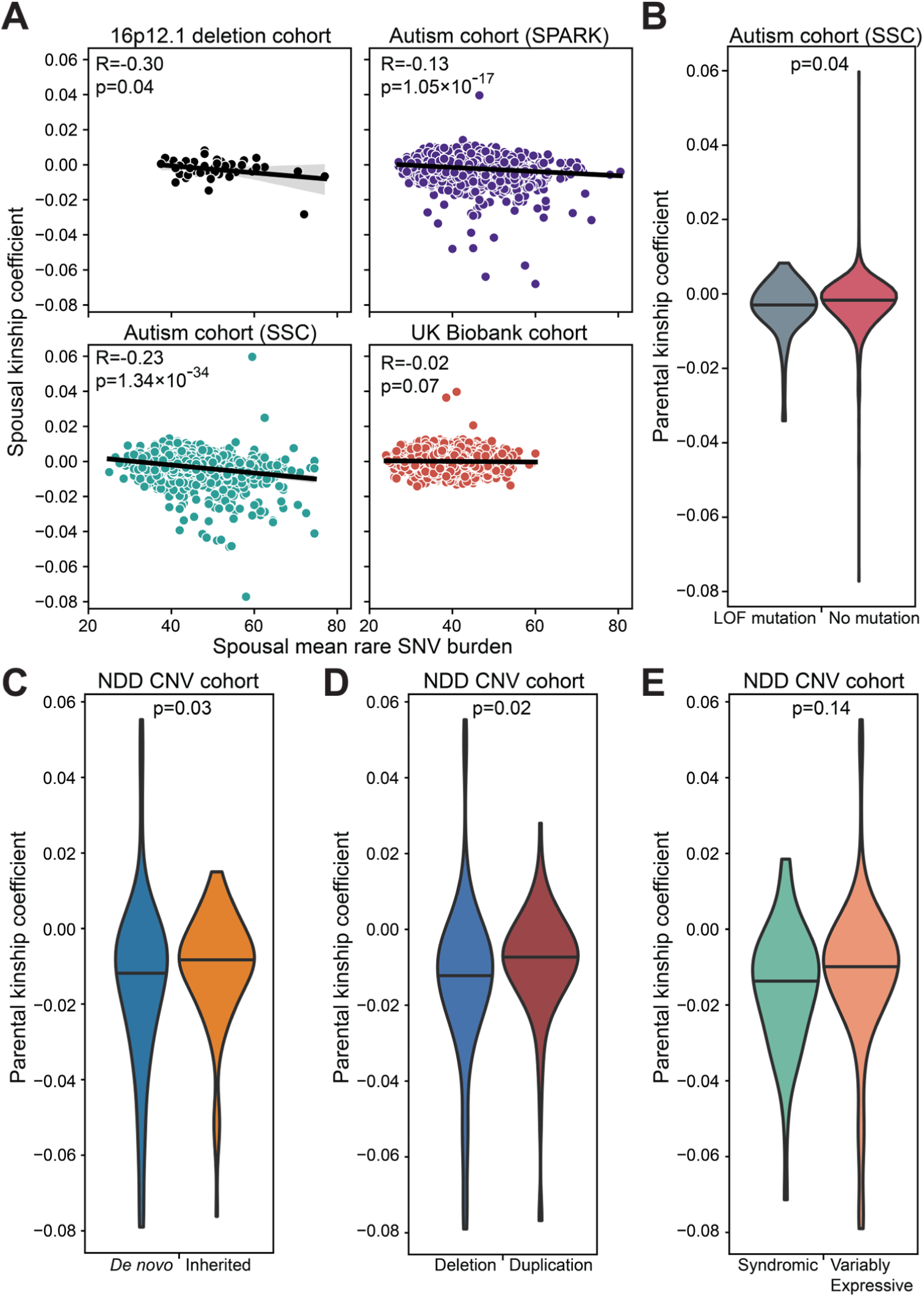
Parental relatedness is a risk factor for neurodevelopmental disorders. (A) Pearson correlations of the average burden of rare, likely deleterious variants and relatedness (measured as kinship coefficients) in spouse pairs from the 16p12.1 deletion (n=44), SPARK (n=4,095) and SSC (n=1,813) autism, and UK Biobank (n=6,315) cohorts. (B) Comparison of parental relatedness among SSC probands with loss-of-function mutations in Tier S SFARI genes (n=102) and without mutations in SFARI genes (n=1,243). Lines indicate median. One tailed t-test. (C-E) Comparisons of parental relatedness among probands ascertained for neurodevelopmental disease with (C) *de novo* (n=129) and inherited (n=116) CNVs, (D) deletions (n=249) and duplications (n=97), and (E) syndromic (n=47) and variably expressive (n=299) CNVs. Lines indicate median. One tailed t-test.

These patterns are in accordance with a liability threshold model (LTM), which posits that an individual will manifest disease once the sum of various independent risk factors exceeds a certain threshold^72^. An inverse relationship between all independent risk factors contributing to disease is implied by the LTM, and has been observed previously when examining additive effects of rare and common variants towards neurodevelopmental disease^1,73, 74^. Thus, the negative relationship between parental kinship and rare SNV burden or presence of highly pathogenic mutations supports parental relatedness as a risk factor for variably expressive pathogenic variants, likely through increased levels of genome-wide homozygosity. This is a particularly compelling hypothesis considering the lack of correlation between spousal kinship and mean spousal rare variant burden seen in the UK Biobank pairs (**Fig. 5A**). Unlike the other cohorts, the pairs in the UK Biobank were not ascertained for children with neurodevelopmental disease and thus may not exist within the same range of liability threshold as the other cohorts.

## DISCUSSION

Assortative mating has previously been hypothesized to explain the high degree of heritability for complex neurodevelopmental and psychiatric disorders^54, 75^. However, the implications of assortative mating have not been explored in the context of rare variably expressive pathogenic variants. Here, we confirm that the well-documented patterns of spousal assortment on neuropsychiatric features^24, 51–58^ hold true for families carrying rare disease-associated variants. It is possible that the observed phenotypic similarity in partners may be caused by mechanisms outside of assortative mating, including partner convergence^76^ or, in disease cohorts, caregiver burden on parents of affected children^77^.

However, we find it unlikely that these mechanisms could account for all observed partner similarity for three key reasons. Firstly, previous studies have indicated that convergence has only a modest effect on spousal similarity^76, 78^. Further, of the 27 participants in the 16p12.1 deletion cohort who indicated the age at depression symptom onset, 70% (19/27) began having symptoms before the birth of their first child. Thus, caregiver burden is an unlikely cause of depression in these individuals and, although these data are limited, we expect similar trends for other disorders. Finally, if spousal phenotypic similarity were the result of convergence or caregiver burden, we would not expect to see a correlation in genetic risk between partners. However, as we do see a strong correlation in rare variant burden, we can conclude that active assortment accounts for much of the observed phenotypic similarity between partners.

Partner assortment on these phenotypes drives increases in genetic risk alleles across generations in affected individuals^75^, similar to the observed patterns of increasing secondary variant burden that are well documented in cohorts of variably expressive CNVs^11, 14^ and are concordant with a “two-hit” mechanism of pathogenicity. Under assortative mating, individuals with similar psychiatric phenotypes but distinct underlying genetic etiologies may preferentially mate and have a child who carries genetic risks from both parents, leading to a compounding of genetic burden over generations. Increased disease liability in children resulting from assortative mating has been previously shown^17, 59^, and the implications for this finding in the context of variably expressive rare variants has been previously hypothesized^79^. Here, our analysis provides more evidence to support the relationship between parental assortment on psychiatric phenotypes and compounding genetic burden towards disease severity.

Unique combinations of rare variants arising from recent mutations have been identified as a major contributor to disease^80, 81^ and assortative mating may drive accumulation of those new combinations. This observation has important implications on the trajectory of individuals within a family carrying a variably expressive variant and provides insights into the genetic “anticipation” observed in these families.

We further highlight the role of spousal relatedness towards complex disorders, showing that spousal kinship acts as a risk factor for variably expressive variants. Long stretches of homozygosity have previously been implicated in neurodevelopmental disorders^68, 69^ and we show here that parental relatedness increases genome-wide homozygosity and, likely through this mechanism, acts as a risk factor for neurodevelopmental disease. Increases in spousal kinship are often due to population stratification^76^, which has heretofore been an underexplored risk factor for neurodevelopmental disorders and may represent an exciting new avenue for further research.

While these results have interesting implications for our understanding of disease pathogenicity, our study has some limitations. We restricted our analysis of phenotypic assortment to only a subset of distinct phenotypes, and while we attempted to control for comorbidities that could inflate correlation estimates, it is likely that some cross-disorder contamination remains. Further, all cohorts assessed here were primarily of European descent and further research is needed to understand how assortment and partner similarity may vary across racial and ethnic groups, particularly for disease-associated rare variants. This may be particularly relevant in the context of isolated populations or those practicing endogamy, who, due to their unique recent genetic history, may have a higher load of both parental relatedness and rare variant burden^82, 83^. Further research is required to examine the genetic architecture in these populations and how these changes in architecture may impact disease risk.

In summary, we provide evidence for assortative mating on neuropsychiatric disorders, particularly in parents of children with neurodevelopmental phenotypes. We show patterns of rare variant burden similarities between spouses and highlight compounding of rare variant burden over generations resulting from phenotypic assortment. We also highlight increased spousal kinship as a risk factor for neurodevelopmental disorders through the unmasking of recessive alleles. Overall, our results highlight the importance of assessing parental history of neuropsychiatric disease and parental relatedness for more accurate assessment of neuropsychiatric disease risk in children and counseling of families carrying variably expressive variants.

## Data Availability

All data produced are available online at NCBI dbGaP study accession phs00245. All code for analysis will be available at GitHub (https://github.com/girirajanlab/Assortative_mating_project).

## ACKNOWLEDGEMENTS

This work was supported by grants T32GM102057 to Miss Corrine Smolen and NS122398 and GM121907 to Dr. Santhosh Girirajan from the National Institutes of Health. Dr. Charles Schwartz was supported, in part, by a grant from the South Carolina Department of Disabilities and Special Needs. Dr. Corrado Romano was supported by the Italian Ministry of Health-Ricerca Corrente 2022.

We are grateful to the families in the 16p12.1 deletion cohort who participated in this study. We are grateful to all the families in SPARK, the SPARK clinical sites and SPARK staff. We are also grateful to all of the families at the participating Simons Simplex Collection (SSC) sites, as well as the principal investigators (A. Beaudet, R. Bernier, J. Constantino, E. Cook, E. Fombonne, D. Geschwind, R. Goin-Kochel, E. Hanson, D. Grice, A. Klin, D. Ledbetter, C. Lord, C. Martin, D. Martin, R. Maxim, J. Miles, O. Ousley, K. Pelphrey, B. Peterson, J. Piggot, C. Saulnier, M. State, W. Stone, J. Sutcliffe, C. Walsh, Z. Warren, E. Wijsman). We appreciate gaining access to genetic and phenotypic data on SFARI Base.

Approved researchers can obtain the SSC and SPARK datasets described in this study by applying at https://base.sfari.org. This research has been conducted using data from UK Biobank, a major biomedical database. More information about UK Biobank is available at https://www.ukbiobank.ac.uk/.

## DATA AND CODE AVAILABILITY

Microarray and genome sequencing data from participants in the 16p12.1 deletion cohort who consented to having their information shared anonymously will be made available at NCBI dbGaP study accession phs00245. Phenotype and genetic data from the SPARK and SSC autism and UK Biobank cohorts through the cohorts’ data portals: SFARI base (https://base.sfari.org) and UK Biobank database (https://www.ukbiobank.ac.uk), respectively. All code for analysis will be available at GitHub (https://github.com/girirajanlab/Assortative_mating_project).

