## Supplemental for "Assortative mating and parental genetic relatedness drive the pathogenicity of variably expressive variants"

### Assortative mating drives pathogenicity of neurodevelopmental disorder-associated variants

#### SUPPLEMENTAL FIGURES

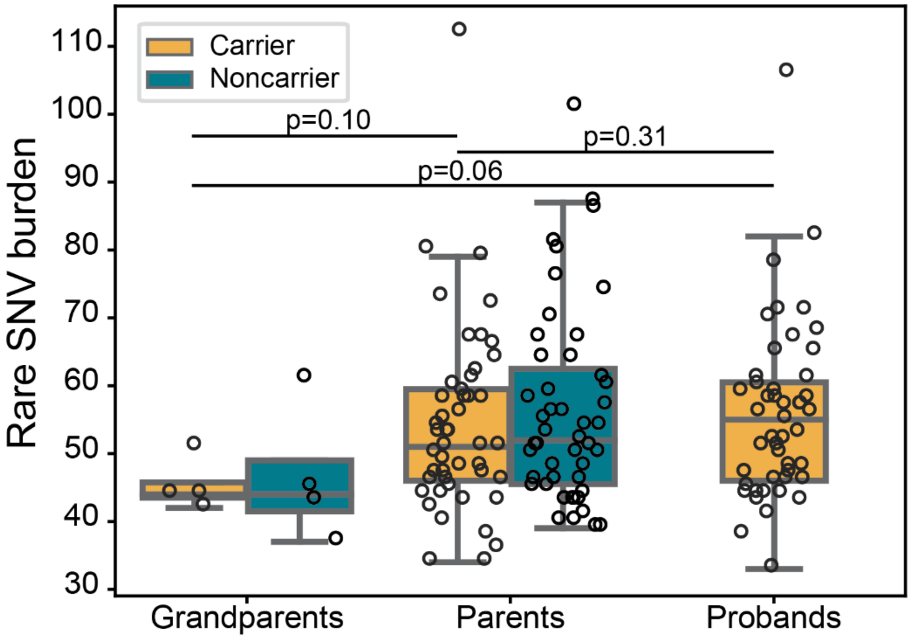

**Figure S1. Increases in rare variant burden in 16p12.1 deletion families.**
Rare, likely deleterious single nucleotide variant (SNV) burden in carrier and noncarrier grandparents and parents and probands in 16p12.1 deletion families. One tailed t-test, comparisons between carriers in each generation.

#### SUPPLEMENTAL TABLES

**Supplementary Table 1. CNVs in the clinically ascertained CNV cohort.**

| **Region** | **Number of families** | |
|---|---|---|
| | **Deletion** | **Duplication** |
| *Syndromic* | | |
| 5q35 (Sotos syndrome region) | 1 | 0 |
| 7q11.23 (Williams syndrome region) | 15 | 7 |
| 15q11-13 (Prader-Willi syndrome region) | 17 | 0 |
| 17q11.2 (Smith-Magenis/Potocki-Lupski) | 3 | 4 |
| 17q21.31 | 3 | 0 |
| **Total Syndromic** | **39** | **11** |

| | | |
|---|---|---|
| *Variably Expressive* | | |
| 1q21.1 | 18 | 26 |
| 3q29 | 6 | 0 |
| 15q11.2 | 32 | 0 |
| 15q13.3 | 19 | 0 |
| 16p13.11 | 20 | 21 |
| 16p11.2 | 63 | 35 |
| 16p11.2 distal | 19 | 4 |
| 16p12.1 | 8 | 0 |
| 17q12 | 11 | 0 |
| 22q11.2 (DiGeorge/VCFS region) | 25 | 0 |
| **Total Variably Expressive** | **221** | **86** |

**Supplementary Table 2. Phenotypes assessed in the Simons Simplex Collection cohort.**

| Phenotype | Reference |
|---|---|
| Autism Diagnostic Observation Schedule (ADOS): social affect | 1 |
| ADOS: restricted and repetitive behavior domain total score | 1 |
| Autism Diagnostic interview (ADI): communication (verbal) domain score | 2 |
| ADI: restricted and repetitive behavior domain total score | 2 |
| ADI: social domain total score | 2 |
| Repetitive Behaviors Scale (RBS) | 3 |
| Parent-reported Social Responsiveness Scale (SRS): total raw scores | 4 |
| Social Communication Questionnaire (SCQ) | 5 |
| Insistence of sameness factor | 6 |
| Social interaction factor | 6 |
| Sensory-motor behavior factor | 6 |
| Self-injurious behavior factor | 6 |
| Idiosyncratic repetitive speech and behavior | 6 |
| Communication skills factor | 6 |
| Vineland Adaptive Behavior Scales: composite standard scores | 7 |
| Full-scale IQ | |
| Verbal IQ | |
| Nonverbal IQ | |
| Developmental Coordination Disorders Questionnaire | 8 |
| Child Behavior Checklist (CBCL) 2-5 internalizing | 9 |
| CBCL 2-5 externalizing | 9 |
| CBCL 6-18 internalizing | 9 |
| CBCL 6-18 externalizing | 9 |

**Supplementary Table 3. Psychiatric phenotype frequencies in SPARK and the UK Biobank.**

| Disorder | SPARK frequency | UK Biobank frequency |
|---|---|---|
| Anxiety | 18.35% (2254/12284) | 6.40% (2943/45958) |
| Bipolar Disorder (BPD) | 3.54% (435/12284) | 0.16% (74/45958) |
| Depression | 21.13% (2596/12284) | 8.92% (4099/45958) |
| Personality Disorder | 1.38% (170/12284) | 0.06% (29/45958) |
| Schizophrenia | 0.47% (58/12284) | 0.03% (12/45958) |
